# Paracoracoid versus Costoclavicular Approach to Infraclavicular Block: A Prospective, Two Arm, Parallel design, Single-centre Randomized Controlled Trial

**DOI:** 10.1101/2020.10.03.20202275

**Authors:** Brigid Brown, Pauline Magsaysay, Yves Bureau, Janice Yu, Shalini Dhir

**Author notes:** Address correspondence to: Shalini Dhir, FRCPC, Department of Anesthesia and Perioperative Medicine, St. Joseph’s Health Care, Western University, 268 Grosvenor’s Street, London, Ontario, Canada, Twitter: @DhirShaliniMD. Conflicts of Interest: The authors declare no conflicts of interest. Funding: The authors have no sources of funding to declare for this manuscript.

## Abstract

**Introduction:** Paracoracoid approach to the brachial plexus block is the conventional infraclavicular technique for upper limb surgeries. In this approach, the ultrasound transducer is placed in the parasagittal plane below the clavicle, medial to the coracoid process. In this view, three cords are separated from each other and are rarely visualized in a single ultrasound window. In the costoclavicular approach, the ultrasound transducer is placed parallel to and below the clavicle. In this view, the cords are clustered together, at a more superficial level. We conducted a randomized controlled trial to compare these two infraclavicular brachial plexus approaches.

**Methods:** Seventy patients were randomized to receive either a paracoracoid or costoclavicular infraclavicular block. Both groups received 35 ml of 0.5% ropivacaine under ultrasound guidance. The primary outcome was sensory block onset time while secondary outcomes included performance times, complications during block insertion (paresthesia, vascular puncture, pleural puncture), block failure, patient satisfaction, and postoperative complications. Telephone follow-up was done 24 h and 7 days later.

**Results:** Sensory block onset time was significantly shorter in the paracoracoid group 18.7 (4.4) min versus 22.2 (6.2) min (p=0.045). Block success at 30 minutes was the same between both groups. There was no difference in any secondary outcomes.

**Conclusions:** This randomized controlled trial demonstrated that the novel costoclavicular approach to the infraclavicular brachial plexus block had similar procedure time, block success and similar complication rates for upper limb surgery when compared to the traditional paracoracoid technique. However, it resulted in longer sensory block onset time.

## INTRODUCTION

The infraclavicular brachial plexus block is a well-recognized regional anesthesia technique for surgeries performed on the distal arm.^1^ In the last fifteen years, the use of ultrasound guidance has become popular for this conventional approach.^2^ Though multiple approaches have been described, paracoracoid approach is traditionally used, worldwide.^3, 4^ In this approach, all three cords of the brachial plexus are separated from one another (figure 1a and 1b) and may not be seen in a single ultrasound window.^5-7^ There can also be significant variation in the position of the individual cords relative to the axillary artery.^6^

**Figure 1a.**
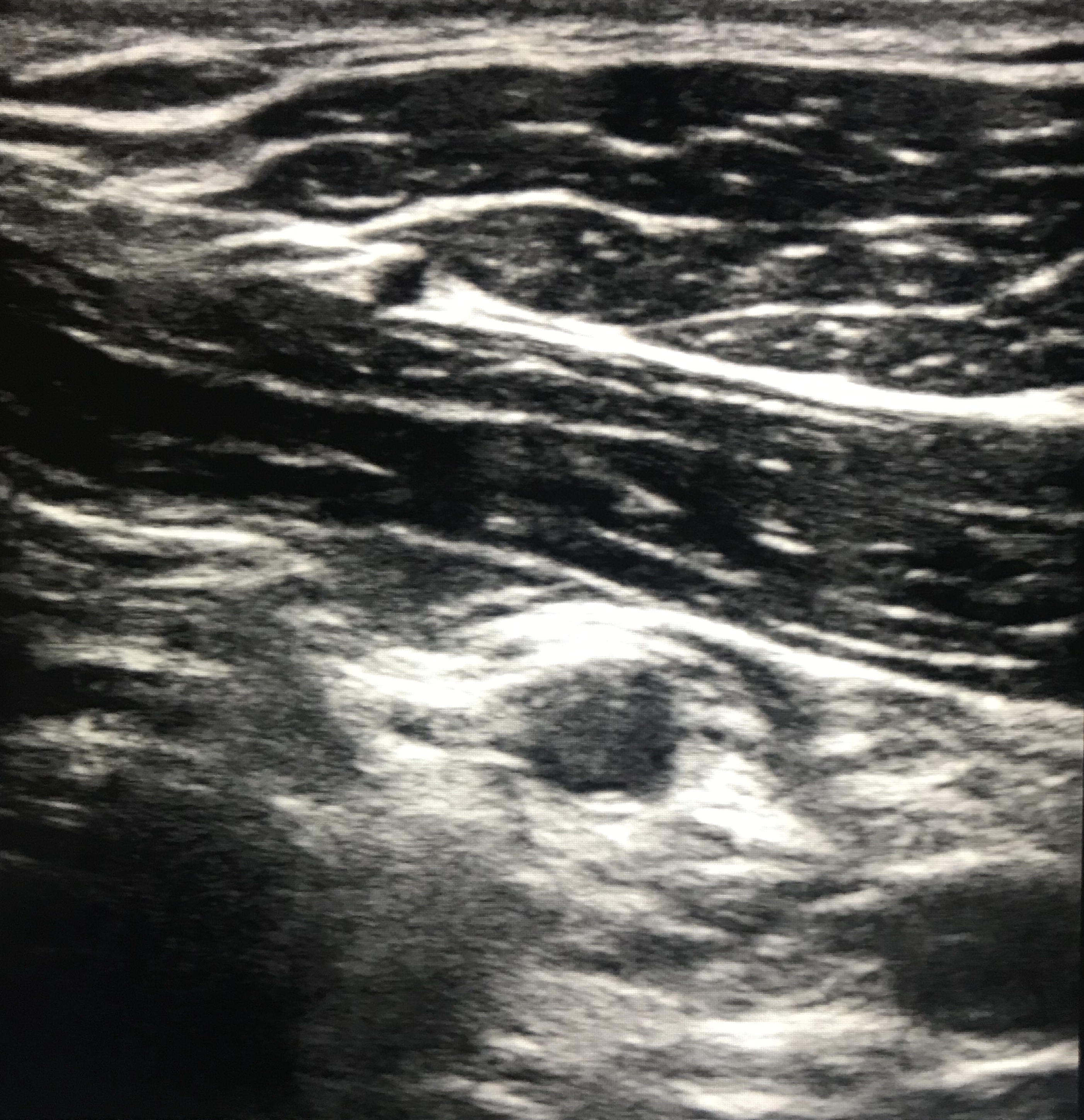
Ultrasound scan as seen by the Paracoracoid approach.

**Figure 1b.**
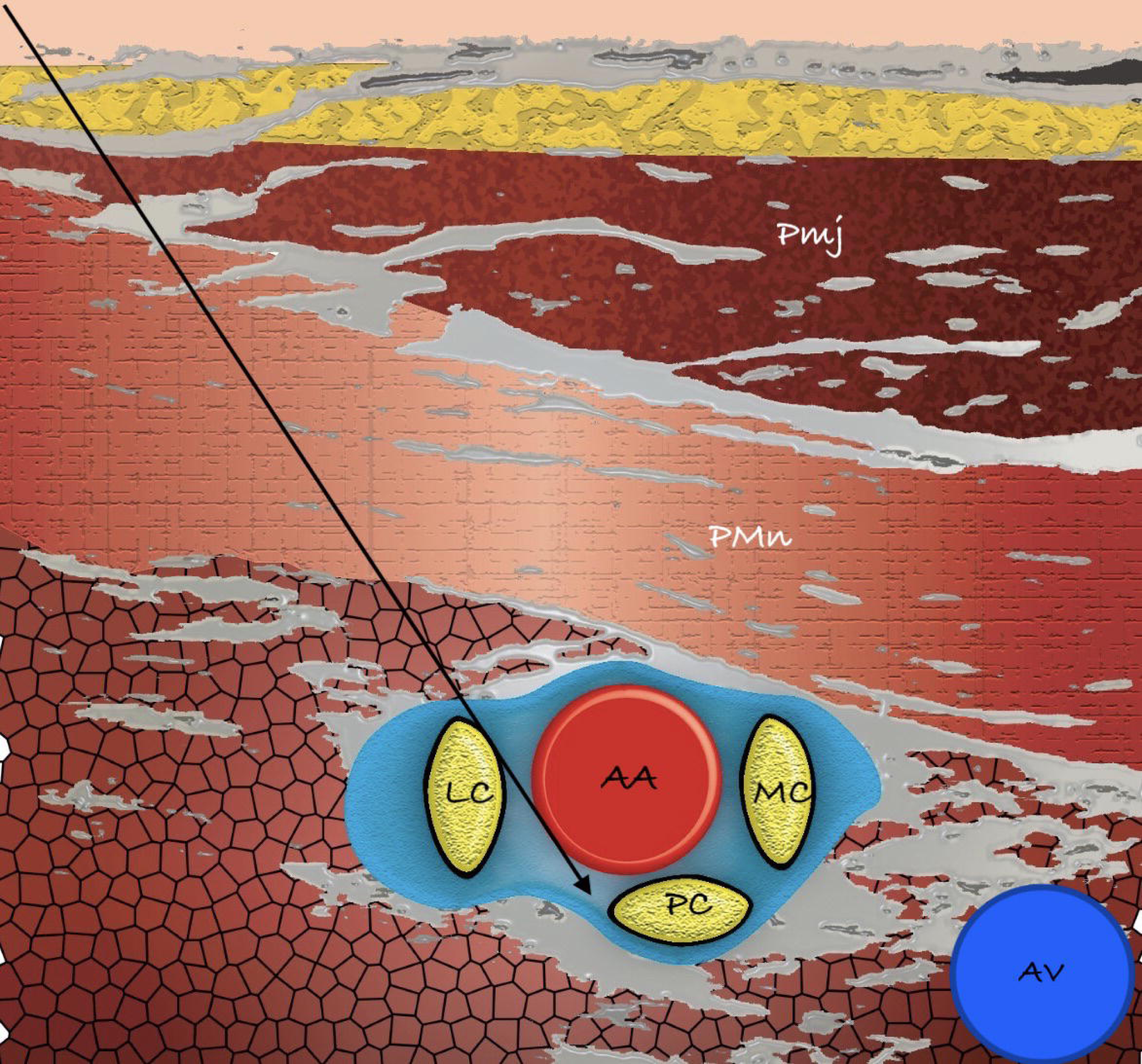
Schematic representation showing the relationship of the cords to the axillary artery and the path taken by the block needle during the Costoclavicular infraclavicular approach to the Brachial Plexus. Abbreviation: AA: axillary artery, LC: lateral cord, MC: medial cord, PC: posterior cord, PMj: Pectoralis major

A new approach to the infraclavicular block has been described in which the ultrasound transducer is placed parallel and inferior to the clavicle and the block needle is inserted in-plane from a lateral to medial direction into the costoclavicular area, the space between the clavicular head of the pectoralis major and the subclavius muscle anterior and the posterior surface of the clavicle and the second rib, posteriorly. In this approach, the cords are clustered together around the lateral edge of the artery at a more superficial level compared with the paracoracoid view (figure 2a and 2b).^8^ Several researchers have suggested that the cords and needle may be easier to visualize with less needle manipulation during the procedure, potentially resulting in a faster onset of the block and a lower incidence of complications.^9-12^

**Figure 2a.**
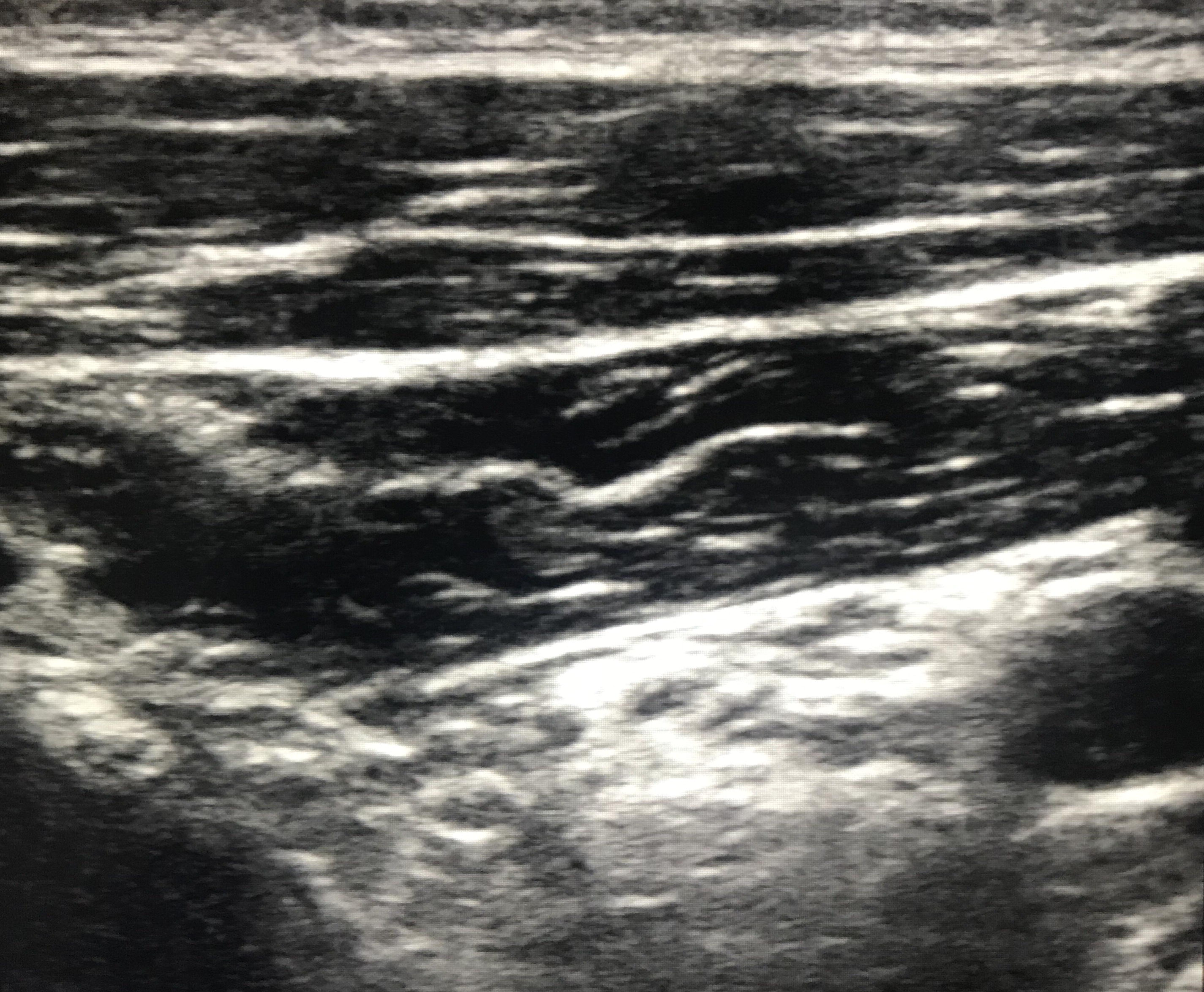
Ultrasound scan as seen by the Costoclavicular approach.

**Figure 2b.**
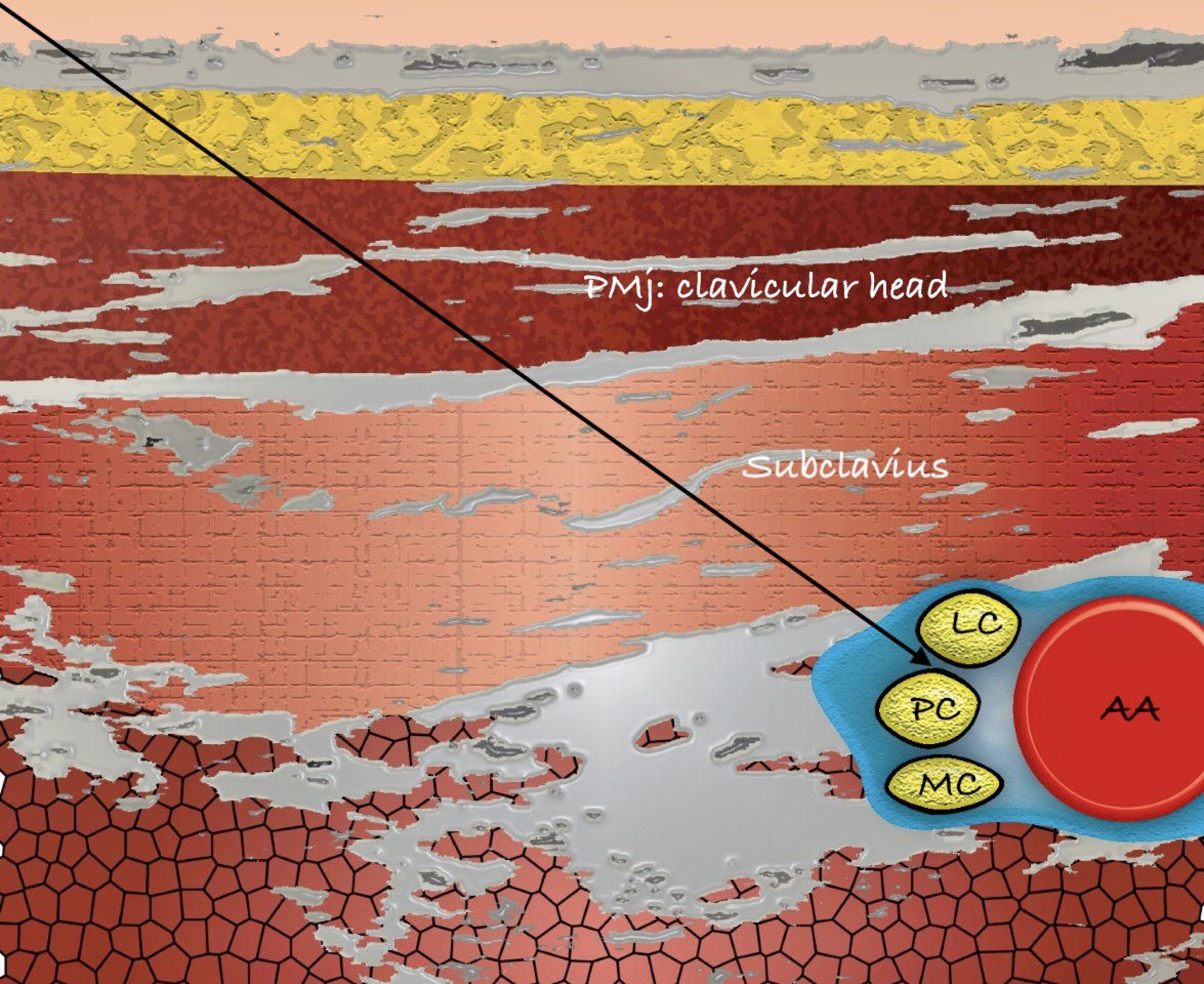
Schematic representation showing the relationship of the cords to the axillary artery and the path taken by the block needle during the Paracoracoid infraclavicular approach to the Brachial Plexus. Abbreviation: AA: axillary artery, AV: axillary vein, LC: lateral cord, MC: medial cord, PC: posterior cord, PMj: Pectoralis major, PMn: Pectoralis minor

The primary aim of this study was to compare the block onset time of the novel costoclavicular (CC) with the conventional paracoracoid (PC) approach to the infraclavicular brachial plexus. We undertook a randomized controlled trial to compare the block onset time with both techniques and to test the hypothesis that the CC approach was similar to the well-established traditional approach. Secondary endpoints included procedure time, procedure related pain and complications, surgical block success, block supplementation and patient satisfaction.

## METHODS

After obtaining institutional ethics approval (#106875, Western University, London, Ontario), the study was registered with clinicaltrials.gov (NCT02657291) on 10 December 2015. After written informed consent, we prospectively enrolled 70 adult ASA I-III patients aged 18-80 years scheduled for elective ambulatory upper limb surgery of the elbow, forearm, wrist, or hand under regional anesthesia. Exclusion criteria included coagulopathy, local anesthetic drug allergy, local site infection or scarring, BMI greater than 35 kg.m^-2^, failure to understand pain scores, pregnant or breastfeeding, and chronic narcotic use. The study was conducted at the Hand and Upper Limb Centre, St. Joseph’s Hospital in London, Ontario between December 2015 and April 2017. Applicable Equator guidelines were followed and the CONSORT 2010 and its extension for reporting multi-arm trial parallel group trial were met.

Using a computer-generated randomization, patients were randomly assigned to one of two groups, each with 35 subjects, to receive an infraclavicular block with either a paracoracoid or costoclavicular approach. Group allocations were concealed in opaque envelopes until opened by the anesthesiologist performing the block. This person had no further involvement in the data collection. All blocks were initiated in a dedicated block room space under aseptic conditions, following intravenous access and the application of standard monitoring and performed by anesthesiologists well versed with both techniques (SD, BB, PM). All blocks were ultrasound guided. Lidocaine 1% (3-5 ml) was used for subcutaneous infiltration prior to initiation of the block, with further supplements as necessary. A Sonosite M-Turbo ultrasound (SonoSite Inc., Bothell, WA, USA) with a high-frequency 10-6 MHz linear array transducer probe with a sterile cover and an 80 mm needle (Pajunk® Geisingen, Germany) were used for all blocks. All patients received a total of 35 ml of 0.5% ropivacaine delivered in aliquots. Patients in both groups also received an intercostobrachial nerve block with 5 ml of 1.5% lidocaine for tourniquet pain.^13^ Complications during block performance such as vascular puncture and paresthesia with needle placement were documented. Once the block was completed, both the possible injection sites were concealed with a dressing to ensure observer blinding.

### Block performance

#### The paracoracoid approach

An ultrasound guided parasagittal paracoracoid (deltopectoral) was used. The needle was inserted in an in-plane approach, at the six o’ clock location, just inferior to the axillary artery.^14^ Aiming at the posterior cord location, two thirds of the drug was deposited at the six o’clock position while the remaining one third was deposited at the nine o’clock position aiming at the lateral cord, during needle retraction.

#### The costoclavicular approach

An ultrasound guided approach as described by Karmakar et al was used.^15^ The ultrasound transducer was placed parallel and inferior to the clavicle and angled cephalad to optimize the ultrasound view. The block needle was inserted in-plane from a lateral to medial direction into the costoclavicular space and the entire drug was deposited in this location. After the block and dressing placement, a blinded observer (JY) unaware of block assignments and not present during block placement performed all assessments of block onset. Motor and sensory block onset of median, radial, ulnar, musculocutaneous, and axillary nerves was evaluated every 5 minutes until the block was complete, or until 30 minutes had elapsed. Sensory block was tested with ice in the appropriate dermatomes of the brachial plexus (2 = normal sensations; 1 = loss of some cold sensations; and 0 = total loss of cold sensations). An effective sensory block was a score of 0 in all nerve distributions. Concurrently, motor block was evaluated (median: thumb opposition; ulnar: finger abduction; radial: wrist/elbow extension; musculocutaneous: elbow flexion) using a similar scale (2 = normal movements possible, 1 = reduced movements but not able to perform movements against resistance, and 0 = total loss of motor power). For incompletely blocked patients at the end of 30 minutes, the surgery proceeded either with supplemental blocks and/or deep sedation or general anesthesia. A single unblocked nerve territory was supplemented distally, at the axilla or the elbow. If there were more than one unblocked nerve, a plexus block was repeated at a different site. If there was insufficient time for local anesthetic supplementation, patients received a general anesthetic.

### Outcomes measurements

The primary outcome for this study was the time to sensory block onset. Secondary outcomes included procedure time, block failure leading to technique supplementation or conversion to general anesthetic (GA), complications during block insertion (vascular puncture, pleural puncture, and paresthesia), other postoperative complications and patient satisfaction. Patients whose block data could not be collected were excluded from the analysis. Patients who required deep sedation because they did not want to be awake/aware during the surgery were not considered block failures and were included.

All patients were telephoned on postoperative day 1 and 7 by the blinded observer who asked standardized questions regarding postoperative complications including nausea, vomiting, opioid use, weakness, persistent paresthesia, pain (standardized scale from 0 = no pain to 10 = worst pain imaginable) and overall block/anesthesia related satisfaction (standardized scale from 0 = completely dissatisfied to 10 = completely satisfied). All adverse events were followed up until resolved.

### Statistical Analysis

The sample size estimation for this study was based on mean onset time of the sensory block. We used comparison of two means to calculate the sample size with a type 1 error of 5% (two-sided alpha of 0.05) and power (1-beta) of 85%.^16^ We determined a priori that the difference of 5 minute was the smallest effect that was acceptable such that a difference of less than 5 minute would be clinically insignificant. Assuming an onset time standard deviation (SD) of 7.1 minutes taken from a previously published study,^17^ we calculated that 58 patients were required to achieve a power of 85% to declare that the lower limit of a one-sided 95% confidence interval (or equivalently a 90% two-sided confidence interval) would be above the limit of -5.^18^ To accommodate for loss of retention, the enrollment was increased to a total of 70 patients for this study. From the same study, we calculated the procedure time SD (appendix 1), and found that this sample size was also sufficient to be 80% sure that the lower limit of a one-sided 95% confidence interval (or equivalent 90% two-sided confidence interval) was above the limit of -12 sec. We did a post-hoc power calculation based on the mean (SD) onset time of 20.8 (6.4) min for all study patients and found that the study was powered to detect a 5 min difference in mean onset time at p = 0.05 and 90% power.

We used IBM SPSS (Ver. 23 SPSS Inc, Chicago Il, 2015 for the statistical analysis. Analysis of all continuous variables (time to perform the block, sensory/motor block onset) was done by using student’s t-test. For dichotomous variables, Pearson’s chi squared test was used. All analyses were considered significant when the resulting type 1 error was less than 0.05 (p<0.05).

## RESULTS

For this study, 74 patients were assessed for eligibility, of which 70 were randomized, with 35 assigned to each group (figure 3 - consort diagram). The final analysis included 30 in PC group and 34 in CC group. One patient in the CC group had local anesthetic systemic toxicity (LAST) and was resuscitated with intralipid. This patient was excluded from analysis. There were 5 patients in the PC group whose assessment could not be completed. The two groups were comparable in terms of patient characteristics and surgical procedures performed (table 1).

**Table 1.** Subject characteristics according to inclusion in primary analysis.

|  | Paracoracoid (PC) | Costoclavicular (CC) |
| --- | --- | --- |
|  | (n=30) | (n=34) |
| **Height cm** | 171.0 (10.9) | 168.7 (9.5) |
| **Age years** | 47.4 (19.2) | 49.4 (15.7) |
| **ASA** |  |  |
| **1** | 10 | 12 |
| **2** | 15 | 18 |
| **3** | 5 | 4 |
| **Male: Female** | 19:11 | 15:19 |
| **Weight kg** | 80.2 (14.2) | 74.5 (13.5) |
| **BMI kgm<sup>-2</sup>** | 27.6 (5.5) | 26.2 (4.6) |
| **Surgery types** |  |  |
| **Elbow** | 6 | 7 |
| **Wrist** | 13 | 14 |
| **Hand** | 11 | 13 |

*Data are presented as mean (SD) or number* *Abbreviations: ASA: American Society of Anesthesiologists classification, BMI: body mass index*

**Figure 3.**
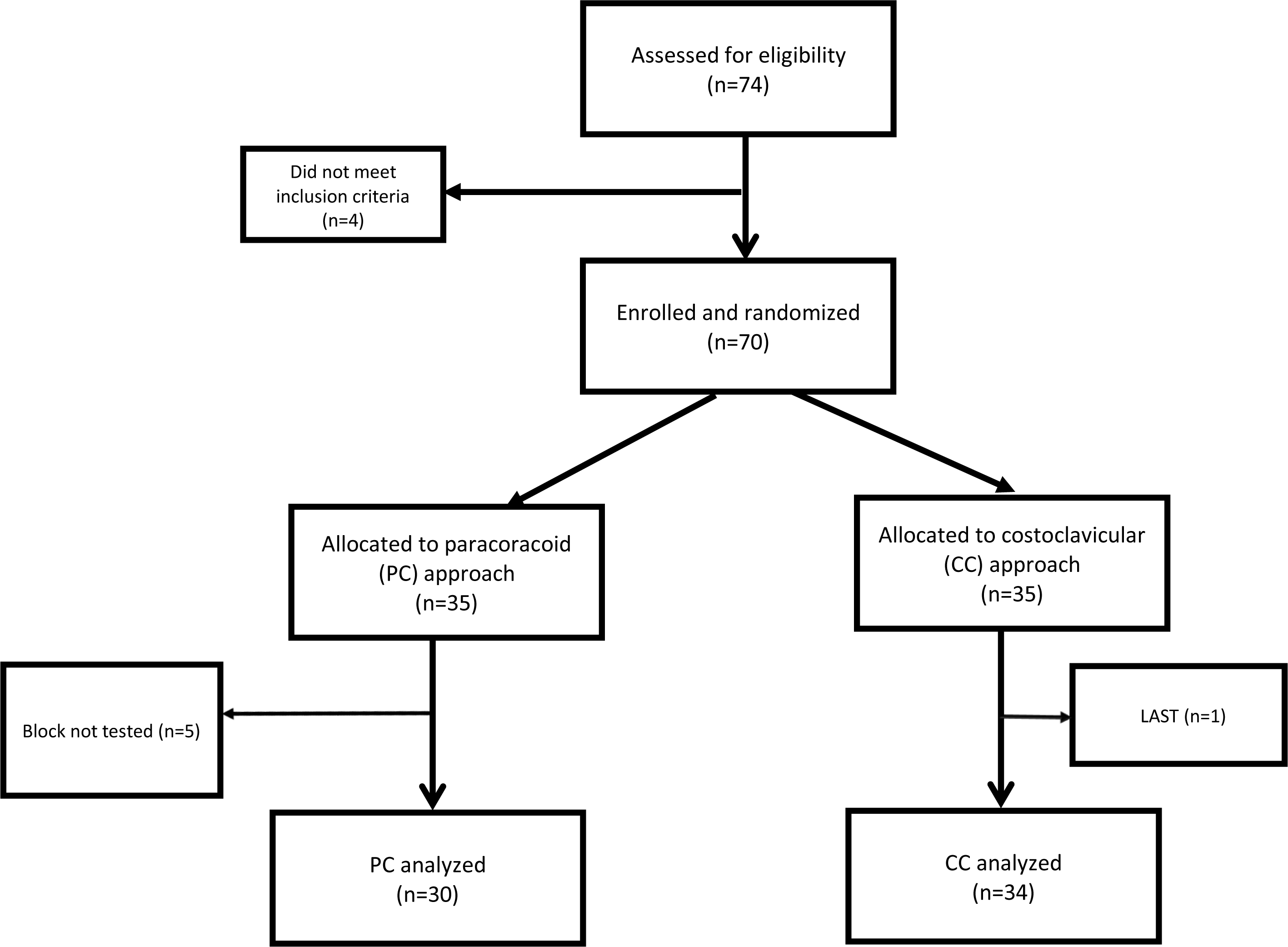
Consort diagram of patient recruitment. Abbreviation: LAST: Local Anesthetic Systemic Toxicity, CC: costoclavicular group, PC: paracoracoid group

The initial imaging time between the groups showed no difference (PC 26.7 versus CC 27.1 sec, p=0.9). The needling time was significantly different between the groups (PC 162.2 versus CC 188.5 sec, p=0.038). The difference in the total procedure time (PC 188.9 (54.2) vs CC 215.6 (56.2) sec, p=0.06) was not significant (figure 4). There were no intergroup differences in block failure, complications during block performance or postoperatively (table 2). The need for conversion to GA (PC 1 vs CC 2, p=0.63), local anesthetic supplementation (PC 0 vs CC 2, p=0.18) or rescue block (PC 1 vs CC 2, p=0.63) were similar between the groups (table 2).

**Table 2.** Block Characteristics - Values are reported as number of subjects (n).

|  | Paracoracoid (PC)<br>N=30 | Costoclavicular (CC)<br>N=34 | p-value |
| --- | --- | --- | --- |
| **Block Interventions** |  |  |  |
| Repeat block | 1 | 2 | 0.63 |
| Surgical supplementation | 0 | 2 | 0.18 |
| GA | 1 | 2 | 0.63 |
| **Procedure related complications** |  |  |  |
| Paresthesia | 3 | 0 | 0.06 |
| Vascular Puncture | 1 | 0 | 0.28 |
| **Postoperative outcomes** |  |  |  |
| Nausea/Vomiting | 2 | 2 | 0.95 |
| Numbness/Tingling | 5 | 6 | 0.92 |
| Weakness | 0 | 2 | 0.16 |
| Satisfaction | 30 | 34 | 0.96 |

**Figure 4.**
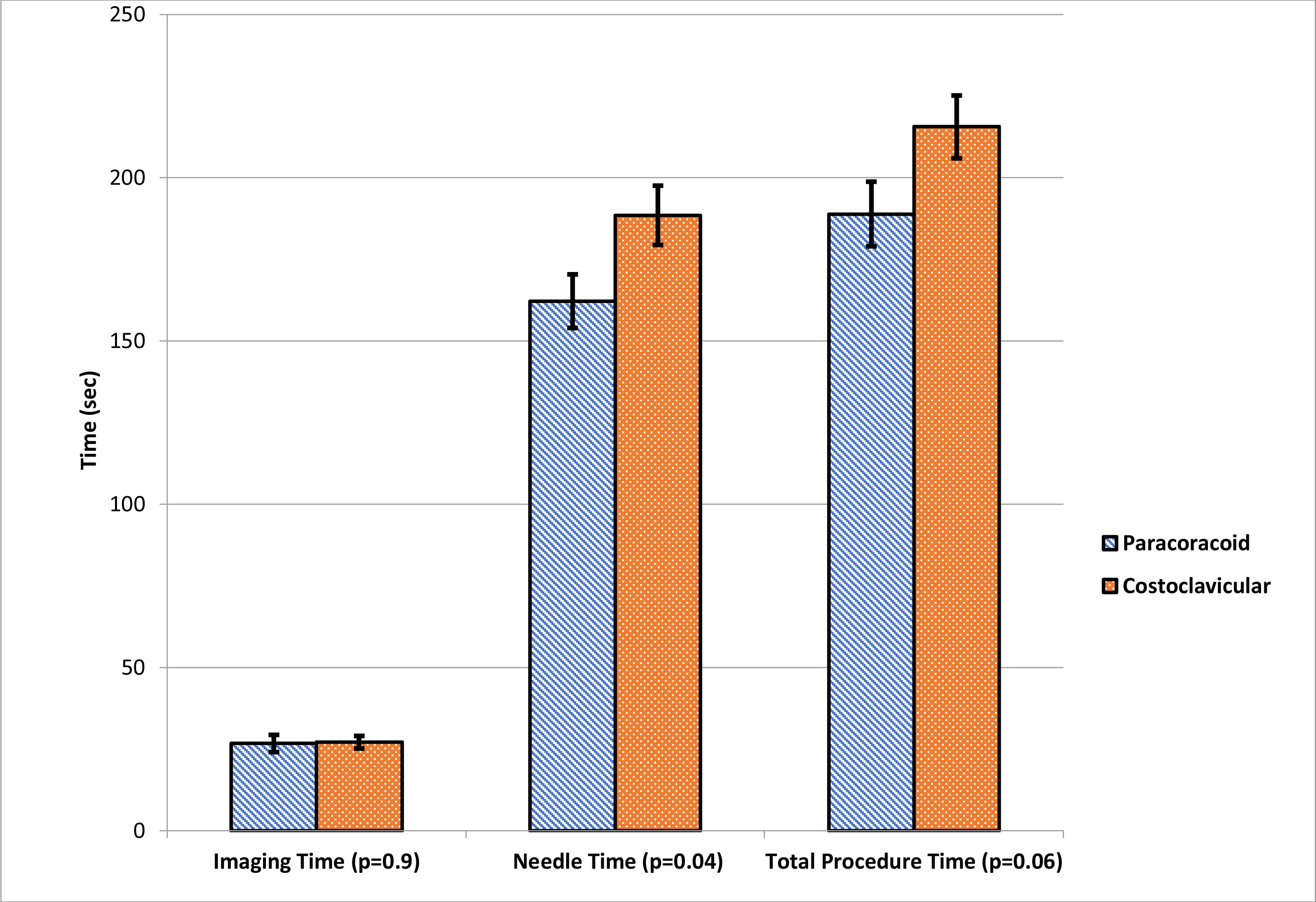
Procedure times. The boxes represent mean time in seconds and the whiskers show SEM.

The primary outcome measure of this study was the sensory block onset. There was a statistically significant faster onset in the paracoracoid approach 18.7 (4.4) min compared with the costoclavicular approach 22.2 (6.2) min (p=0.045) (figure 5). Sensory and motor block onset for each nerve in the plexus are in table 3. At the end of 30 minutes, complete sensory block co-relating with surgical readiness was 93.3% in PC group and 91.2% in CC group (p=0.75) (figure 6). The block onset pattern differed between the groups over the 30 minutes. The CC group had a faster initial onset with 5 patients (15%) demonstrating a sensory block after 5 minutes compared with only 1 patient (3%) in the PC group. However, this order changed and at 20 minutes, PC group had a higher number of complete sensory block (24 vs 19, p=0.04). Beyond 20 minutes, the sensory block onset was comparable between the two groups. There was no difference in motor block onset (appendix 2).

**Table 3.** Block onset data.

|  | **Paracoracoid**<br>**(PC)**<br>**N=30** | **Costoclavicular**<br>**(CC)**<br>**N=34** | **Mean**<br>**difference** | **95% CI of**<br>**mean**<br>**difference** | **P value** |
| --- | --- | --- | --- | --- | --- |
| **Total sensory**<br>**(min)** | 18.7 (4.4) | 22.2 (6.2) | -1.69 | -6.99, -0.82 | **0.045\*** |
| **Axillary**<br>**(min)** | 15.3 (6.7) | 15.8 (7.5) | 0.50 | -4.01, 5.01 | 0.82 |
| **Median**<br>**(min)** | 12.7(5.9) | 13.3 (5.9) | 1.14 | -2.5, 4.79 | 0.54 |
| **MCT**<br>**(min)** | 11.7 (3.6) | 12.8 (5.5) | 0.29 | -2.9, 3.47 | 0.86 |
| **Radial**<br>**(min)** | 12.7 (3.2) | 13.1 (7.3) | 0.58 | -2.9, 4.09 | 0.74 |
| **Ulnar**<br>**(min)** | 12.7 (5.6) | 17.2 (7.5) | -1.67 | -5.68, 2.34 | 0.41 |
| **Total motor**<br>**(min)** | 20.3 (6.4) | 21.7 (6.7) | -1.41 | -5.81, 3.00 | 0.53 |
| **Axillary**<br>**(min)** | 18.3 (6.7) | 18.1 (7.5) | 0.98 | -3.27, 5.34 | 0.64 |
| **Median**<br>**(min)** | 16.3 (6.4) | 17.2 (7.1) | -1.55 | -5.49, 2.40 | 0.44 |
| **MCT**<br>**(min)** | 16.7 (6.2) | 14.7 (8.5) | 0.95 | -3.82, 4.01 | 0.96 |
| **Radial**<br>**(min)** | 18.7 (7.2) | 15.3 (6.5) | 3.19 | -0.47, 6.85 | 0.86 |
| **Ulnar**<br>**(min)** | 15.0 (6.5) | 15.0 (5.4) | 1.78 | -1.89, 5.46 | 0.34 |
| *Data are presented as mean (SD)* |  |  |  |  |  |
| *\*shows significant difference* |  |  |  |  |  |
| *Abbreviations: MCT: musculocutaneous, CI: confidence interval, Min: minutes* |  |  |  |  |  |

**Figure 5.**
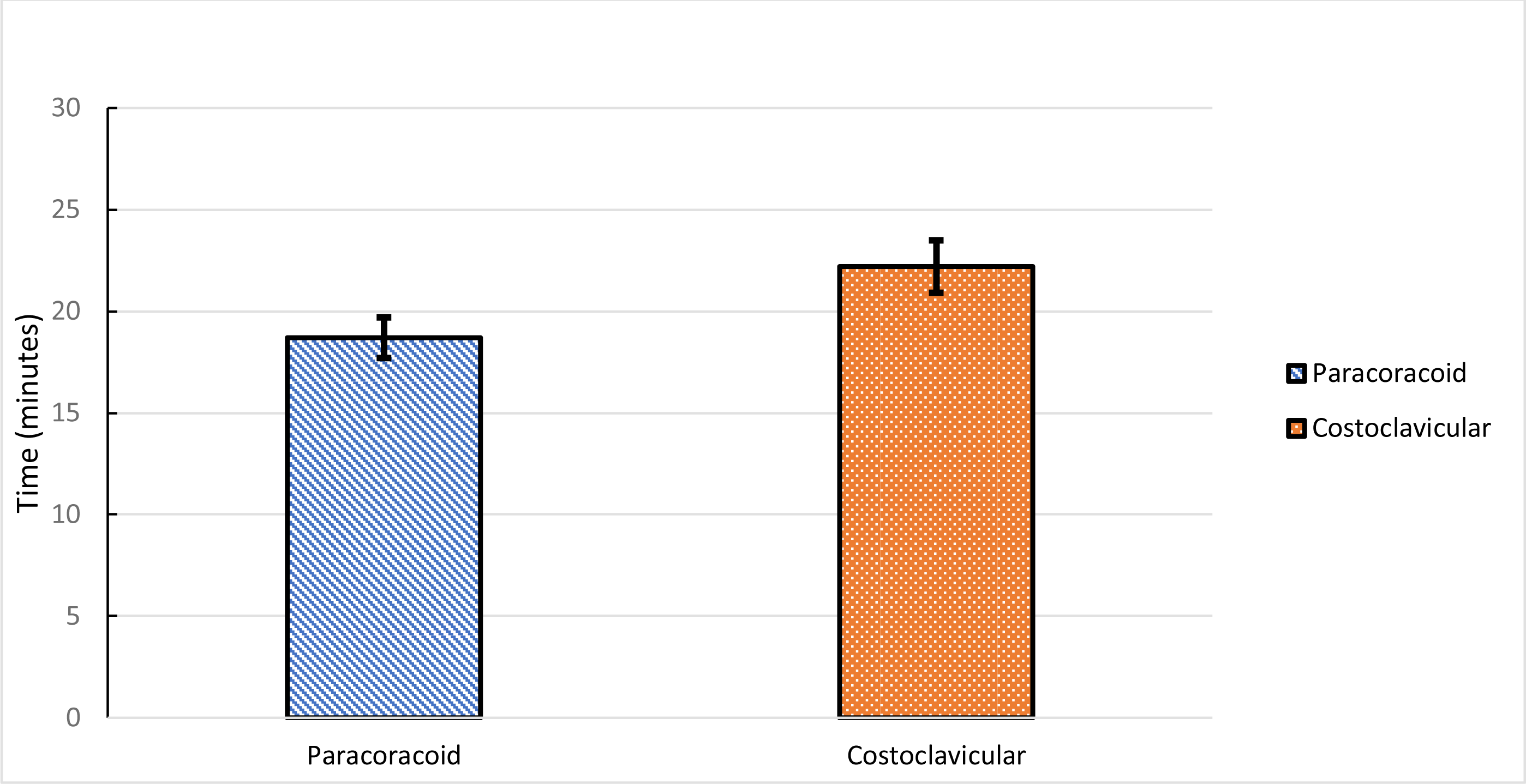
Primary outcome: sensory block onset. The boxes represent mean time in minutes and the whiskers show SEM. The difference was significant (p=0.045)

**Figure 6.**
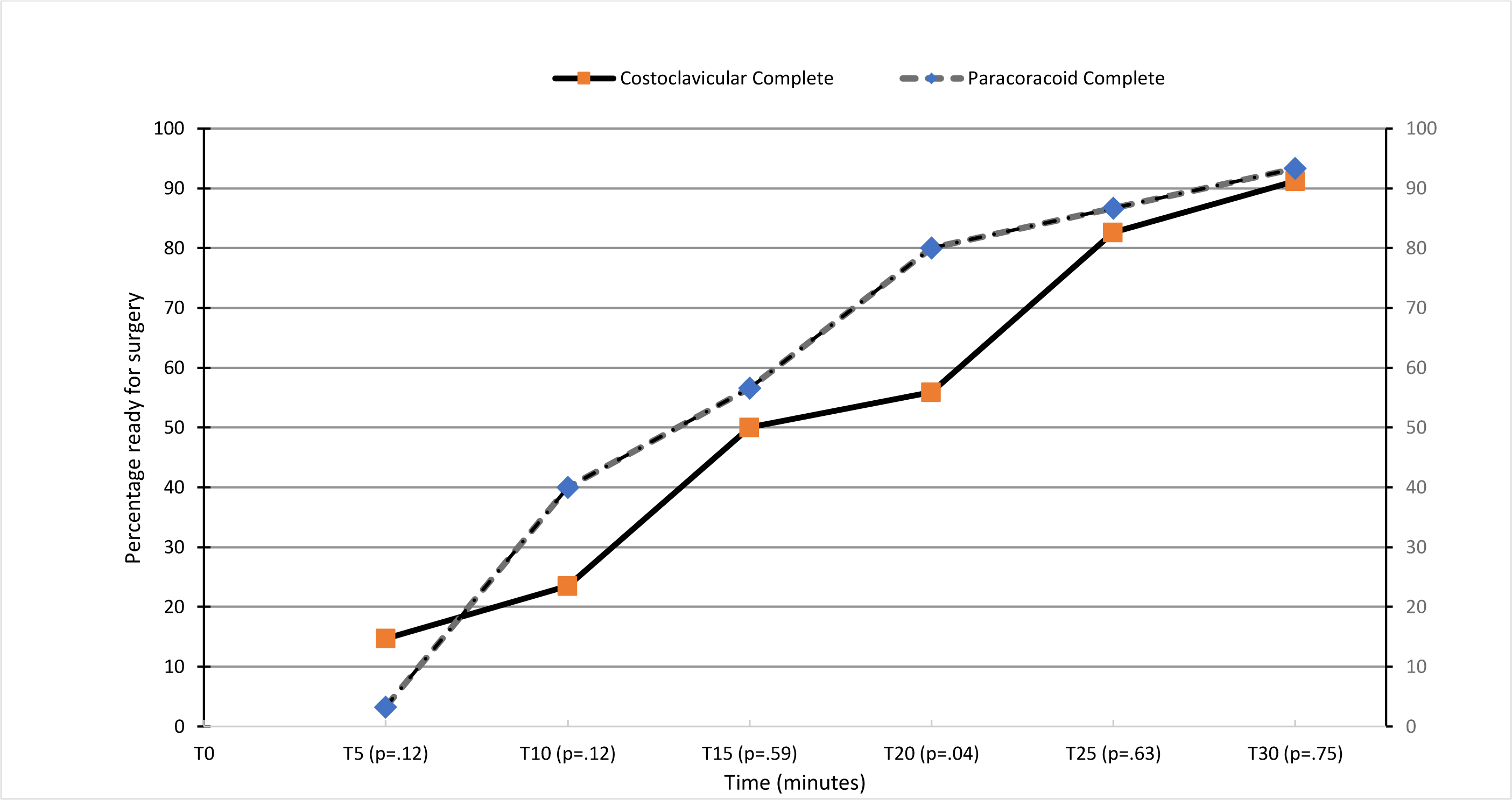
Percentage ready for surgery at various time points during the 30 minutes assessment period.

## DISCUSSION

This observer-blinded, parallel group, two arm randomized controlled study demonstrates that the paracoracoid approach to the infraclavicular brachial plexus block was associated with significantly shorter block onset time when compared to the costoclavicular approach. However, there was no difference in the surgical readiness at the 30 min time point (PC 93.3% versus CC 91.2%, p= 0.75, figure 5). Both techniques had similar ultrasound scanning time. Though the costoclavicular approach took longer needling time and therefore longer overall performance time, the difference missed statistical significance (188.9 versus 215.6 sec, p=0.06, figure 3). Shorter procedure and block onset times are important particularly when the block is used as the primary anesthetic technique. However, this study found no significant difference between the groups.

An interesting finding from this investigation was the pattern of the sensory block onset. In the first 5 minutes after the block performance, the CC group showed sensory block onset in 15% patients whereas only 3% were blocked in the PC group. At 20 min, the difference in the number of blocked patients was significantly different but this difference evened out (figure 5). This pattern was not observed in the motor block onset (figure 6). It is difficult to speculate the reason for this. Previous authors have noted and explained this difference as a result of using large volumes of local anesthetic affecting the pattern of block onset.^19^ While it is possible that the volume theory may account for some component of the block onset, it does not seem to fully explain this phenomenon as the volume used was similar in both groups, consistent with prior studies and do not fully explain the sensory block onset behaviour.^20, 21^ The significance of this sensory block pattern may not be clinically relevant but warrants further studies.

One patient in the CC group developed LAST and required intralipid for resuscitation. Though rare, the quoted incidence of LAST is 0.27% ^22^ and its possibility cannot be excluded, despite very careful technique.

We need to remark on several limitations of this study. First, it was designed as a non-inferiority study. With an equivalence trial on the subject showing no intergroup differences,^19^ we chose a non-inferiority design. It is known that equivalence dismisses the option that the new treatment could be superior. In a non-inferiority design, the difficulty is in choosing the value of delta. The non-inferiority margin or delta of 5 min was based purely on clinical grounds as we had to choose a cut-off point. Second, we chose to exclude protocol violators and did not analyze them as intention-to-treat (ITT). ITT is the recommended method in superiority trials.^23^ It is known that ITT may not have a conservative approach and dropouts may direct the results of the study arms towards each other though per protocol analysis may lead to bias.^24^ On the other hand, both types of analyses support non-inferiority. Third, the sample size chosen was conservative. Though it matched prior studies and was appropriately calculated, modest sample sizes may not lead to conclusive results. Regardless of the sample size, it may not be possible to ever prove that the two techniques are truly identical in their efficacy unless the confidence interval centers on zero or with zero width.^23^ Fourth, we did not target the cords individually but elected to follow the original recommendations of block performance.^15^ It is possible that targeting the cords may have resulted in a faster onset. Fifth, we chose a volume of 35 ml for both techniques. This volume has been established as the minimum effective volume (MEV) for the conventional paracoracoid approach.^20^ Though the appropriate volume for the costoclavicular approach has not been established, a similar MEV has been suggested.^21^ Last, we chose a scoring system of 2-0 for sensory and motor block onset. Many researchers are now using a composite score combining all the nerves and achievement of 70-80% of the score is considered success (Karmakar, M. personal communication with BB, May 2018). We may have been too rigid in choosing a score of 0 (denoting a complete block) in all 5 nerve dermatomes as success.

There has been much interest in a more medial approach to the brachial plexus in the infraclavicular area. Though the costoclavicular approach has been a recent nomenclature, proximal, medial, vertical approaches essentially describe the same location and there have been multiple studies evaluating the block characteristics of these approaches.^9, 11, 15, 25^ Results from past studies comparing the two techniques have been equivocal. Some technical reports have led us to believe that medial approaches may be better and quicker for infraclavicular block.^8, 9, 19, 26^ Multiple studies evaluating the feasibility of the costoclavicular approach to the infraclavicular block have been undertaken but there are very few studies directly comparing the two approaches albeit with different results. Leurcharusmee et al compared the costoclavicular approach to the conventional paracoracoid approach and found no difference.^19^ The study by Mosaffa and colleagues compared the nerve stimulation guided conventional coracoid approach to the more medial vertical infraclavicular approach and found the former to be superior.^27^ Songthamwat et al found faster block onset and readiness for surgery with the costoclavicular approach.^28^ In the present study, we found faster sensory block onset with the conventional paracoracoid approach. The procedure time was not different statistically though there was a mean difference of a few seconds between the two techniques. Both techniques were found to be effective for ambulatory upper limb surgery.

Notwithstanding the results of this study, there may be indications for using the proximal technique for catheter insertion and in obese patients as the space is narrower and shallower in this approach. It is known that the space narrows further with arm abduction and this may be useful in obese patients whose plexus is located deeper in the more distal paracoracoid area.^29^ This may justify the need for further studies to evaluate the feasibility of the costoclavicular approach in special subgroup of patients as well as indwelling regional catheter techniques.

## Conclusion

When compared with the traditional paracoracoid technique, the novel costoclavicular approach to the infraclavicular brachial plexus block resulted in longer sensory block onset time but demonstrated similar procedure time, block success and complication rates for ambulatory upper limb surgery.

## Supporting information

Supplemental table for motor block

## Data Availability

Raw data is available from the corresponding author but is not available as a direct URL.

## ACKNOWLEDGEMENTS

This work was presented at

1. 36th Annual European Society of Regional Anaesthesia & Pain Therapy (ESRA) Congress 2017: ESRA7-0309.
2. Australian and New Zealand College of Anaesthetists Annual Scientific Meeting 2018: accepted for Gilbert Brown prize: abstract #2515.

