## Appendix1 for "Paracoracoid versus Costoclavicular Approach to Infraclavicular Block: A Prospective, Two Arm, Parallel design, Single-centre Randomized Controlled Trial"

Appendix 1.

IQR was converted to standard deviation considering the data to be symmetrically distributed. We used the estimator estimating the standard deviation from IQR is provided in the Cochrane Handbook, which is defined as

𝑆D≈𝑞3−𝑞1/1.35.

The following references were used:

1. Keller T. Re: Is there any way to get mean and sd from median and iqr (interquartile range)? Available at: [https://www.researchgate.net/post/Is_there_any_way_to_get_mean_and_SD_from_median_and_IQR_interquartile_range/53296a11d11b8bce568b45f7/citation/download. Accessed 16 September 2020](https://www.researchgate.net/post/Is_there_any_way_to_get_mean_and_SD_from_median_and_IQR_interquartile_range/53296a11d11b8bce568b45f7/citation/download.%20Accessed%2016%20September%202020).
2. <https://handbook-5-1.cochrane.org/chapter_7/7_7_3_5_mediansand_interquartile_ranges.htm#:~:text=In%20other%20situations%2C%20and%20especially,the%20outcomes%20distribution%20is%20skewed>.
3. Higgins JPT, Green S: Cochrane Handbook for Systematic Reviews of Interventions. 2008, Wiley Online Library.
