## Supplementary figures and images for "Paracoracoid versus Costoclavicular Approach to Infraclavicular Block: A Prospective, Two Arm, Parallel design, Single-centre Randomized Controlled Trial"

### Supplemental table for motor block

Appendix 2:
Motor block onset time at various time points during the 30 minutes assessment period

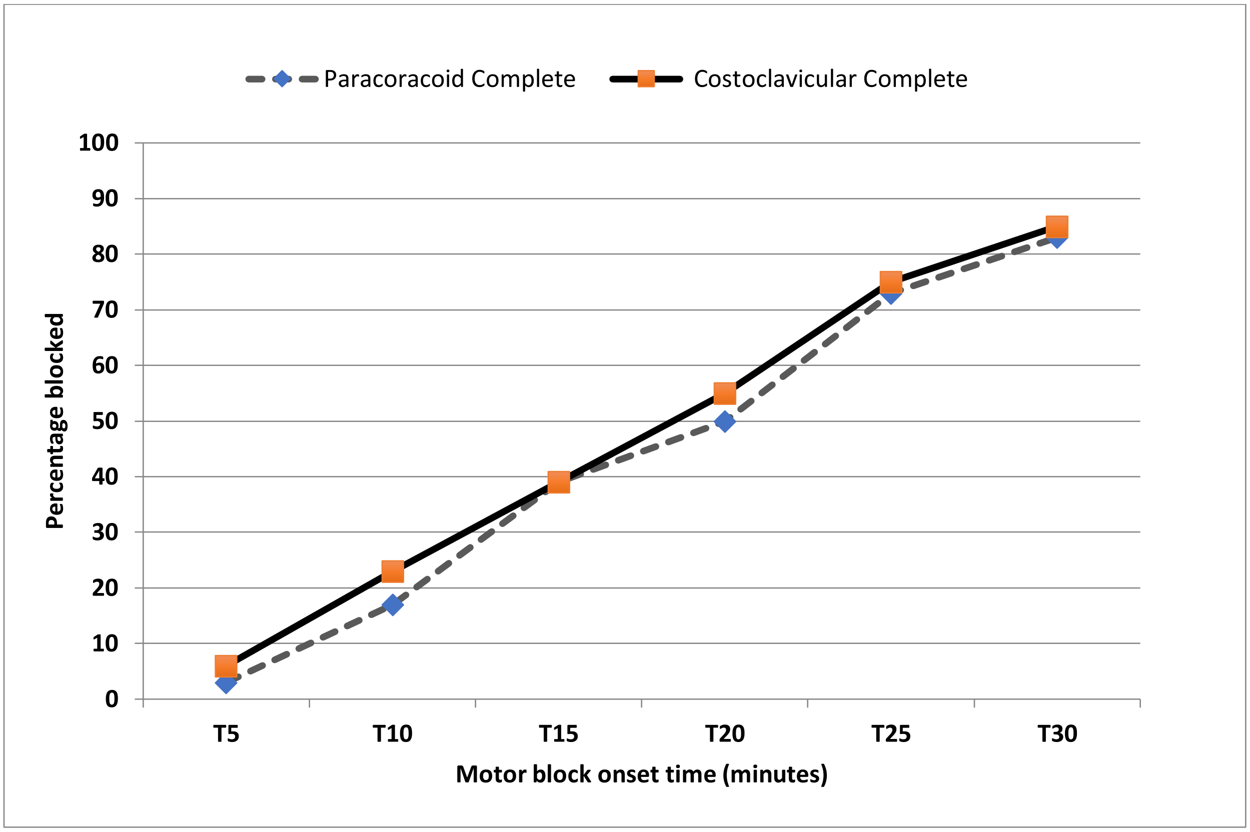
